# Effectiveness of remdesivir with and without dexamethasone in hospitalized patients with COVID-19

**DOI:** 10.1101/2020.11.19.20234153

**Authors:** Brian T. Garibaldi, Kunbo Wang, Matthew L. Robinson, Scott L. Zeger, Karen Bandeen Roche, Mei-Cheng Wang, G. Caleb Alexander, Amita Gupta, Robert Bollinger, Yanxun Xu

## Abstract

**Rationale:** Remdesivir and dexamethasone reduced the severity of COVID-19 in clinical trials. However, their individual or combined effectiveness in clinical practice remains unknown.

**Objectives:** To examine the effectiveness of remdesivir with or without dexamethasone.

**Methods:** We conducted a multicenter, retrospective cohort study between March 4 and August 29, 2020. Eligible COVID cases were hospitalized patients treated with remdesivir with or without dexamethasone. We applied a Cox proportional hazards model with propensity score matching to estimate the effect of these treatments on clinical improvement by 28 days (discharge or a 2-point decrease in WHO severity score) and 28-day mortality.

**Measurements and Main Results:** Of 2485 COVID-19 patients admitted between March 4 and August 29, 2020, 342 received remdesivir and 157 received remdesivir plus dexamethasone. Median age was 60 years; 45% were female; 81% were non-white. Remdesivir recipients on room air or nasal cannula oxygen had a faster time to clinical improvement (median 5.0 days [IQR 4.0, 8.0], remdesivir vs. 7.0 days [IQR 5.0, 12.0], control; adjusted hazard ratio (aHR) 1.55 [1.28; 1.87]), yet those requiring higher levels of respiratory support did not benefit. Remdesivir recipients had lower, but statistically insignificant, 28-day mortality (7.6% [23 deaths], remdesivir vs. 14.9% [45 deaths], control). Adding dexamethasone trended toward lower 28-day mortality compared to remdesivir alone (5.1% [8 deaths] vs. 9.2% [17 deaths]; aHR 0.14 [0.02; 1.03]).

**Conclusions:** Remdesivir offered a significantly faster time to clinical improvement among a cohort of predominantly non-white patients hospitalized with COVID-19, particularly with mild-moderate disease. Remdesivir plus dexamethasone may reduce mortality.

## INTRODUCTION

The pandemic caused by SARS-CoV-2 continues to progress with over 25 million global cases and 850,000 deaths as of September 1, 2020.^1^ While the world awaits an effective vaccine, a number of pharmacologic agents have been studied for COVID-19, the syndrome caused by SARS-CoV-2. Remdesivir, a nucleotide analog prodrug with in vitro effects against a broad array of RNA viruses,^2-4^ has received considerable attention.^3,5,6^ Clinical trials assessing the effectiveness of remdesivir have yielded conflicting results,^7-11^ have not included a sufficient number of patients from groups most affected by COVID-19, such as Black and Latinx individuals,^12^ and have not examined the co-administration of remdesivir with other drugs.

In addition to uncertainty about efficacy, there are concerns that the demand for remdesivir will outpace supply, raising calls for a transparent and just distribution process.^13^ Black and Latinx individuals have mortality rates that are 2.4 and 1.5 times higher than white individuals, respectively.^14^ However, only 11-20% of participants in remdesivir clinical trials were Black, and only 17-23% were Latinx.^8,10,11^ Since underrepresented minorities shoulder a disproportionate burden of morbidity and mortality from COVID-19,^15^ it is critical that we understand the effects of remdesivir in these populations.

The optimal treatment duration of remdesivir also remains unclear. The Adaptive COVID-19 Treatment Trial (ACTT-1) did not find a significant difference in outcomes among patients randomized to a 5-day vs. 10-day course of remdesivir.^10^ In contrast, a recent open label study found that a 5-day treatment course, but not a 10-day course, led to statistically significant improvement at day 11.^11^

Notably, the UK-based RECOVERY trial showed that dexamethasone administration compared to placebo led to a significant reduction in mortality (22.9% vs. 25.7%).^16^ Whether there is additional benefit from using both remdesivir and dexamethasone requires further evaluation. We used time-dependent propensity score matching to compare the effectiveness of remdesivir to routine care among patients admitted to our 5-hospital health system from March 4 through August 29, 2020.

## METHODS

### Study Design and Participants

This study was conducted at five hospitals (Johns Hopkins Hospital, Baltimore, MD; Bayview Hospital, Baltimore, MD; Howard County General Hospital, Columbia, MD; Suburban Hospital, Bethesda, MD; Sibley Hospital, Washington DC) that comprise the Johns Hopkins Medicine System (JHM) with 2,513 beds serving approximately 7 million persons. The hospital institutional review boards approved this study as minimal risk and waived informed consent requirements. All patients consecutively admitted with confirmed SARS-CoV-2 infection by microbiological testing between March 4 and August 29, 2020 were eligible.

### Data Collection

The primary data source was JH-CROWN: The COVID-19 PMAP Registry, which utilizes the Hopkins Precision Medicine Analytics Platform.^17,18^ Some patients were included in a previous description of the cohort.^18^

### Eligibility Criteria and Administration of Remdesivir

The health system remdesivir policy followed the guidance from the Emergency Use Authorization (EUA). Patients prescribed remdesivir were required to have significant illness (SpO2 ≤ 94% on room air or require supplemental oxygen, mechanical ventilation, or extracorporeal membrane oxygenation [ECMO]) and have an alanine aminotransferase level (ALT) less than five times the upper limit of normal. A lottery system was developed in the event of drug shortage, but was not needed during the study period. Patients who were intubated at the end of a 5-day treatment could receive an additional 5-days if they had not improved.

Treatment was stopped if the ALT rose above 5 times the upper limit of normal, or if there were other signs or symptoms of hepatoxicity. Physicians were encouraged to stop remdesivir if the creatinine clearance dropped by more than 50% with no alternative explanation. The decision to prescribe remdesivir was made by the attending physician in conjunction with the patient or their surrogate. Remdesivir requests were approved by the central pharmacy committee. Patients who received at least one dose of remdesivir outside of a clinical trial were defined as remdesivir patients.

### Outcomes

The primary outcome of interest was time to clinical improvement from the start of remdesivir, a composite outcome defined as discharged alive from the hospital without worsening of World Health Organization (WHO) disease severity score or at least a two-point decrease in WHO severity score during hospitalization within 28 days or maximum follow up.^19^ Failure of clinical improvement was censored at the last day of follow up or 28 days, whichever came first. The death outcome was also censored at 28 days.^19^ The secondary outcome was time to death from the first remdesivir treatment day. Patients who were discharged alive were censored at 28 days to account for death and discharge being competing risks. An additional secondary outcome was the combined effectiveness of dexamethasone and remdesivir.

### Statistical Analysis

Cox proportional-hazards regression models were applied to estimate the association between remdesivir treatment and outcomes of interest. A set of demographic, clinical variables and laboratory results were included in Cox regression models based on clinical interest and knowledge. To account for the non-randomized assignments of remdesivir and different timings of initial administration, we used time-dependent propensity score matching to create pairs of individuals, one treated and the other being the most similar patient eligible for treatment at the time of initiation. Analyses were then performed on the matched sets.^20,21^ Patients were included in matching if their admission dates were later than the earliest admission date (April 27, 2020) among patients in the remdesivir group. In addition, a time constraint was imposed so that a patient in the remdesivir group with *k* days of treatment was matched to a patient in the control group who stayed in the hospital at least *k* days (5 days maximum) beyond the matching day.

The last condition avoided matches in which the control patient was healthy enough to be discharged soon after the matched day, as those patients would not be similar to cases who were initiating remdesivir (**see Supplement** for details of matching procedures, variable selections, and missing value imputations). Data was analyzed using R version 3.6.2.

### Sensitivity Analyses

The above analyses were repeated using the same propensity score matching method, but we excluded remdesivir patients who also received dexamethasone and applied ACTT-1^8^ inclusion/exclusion criteria to select qualified patients for matching. We varied matching requirements regarding the number of days matched patients had to remain in the hospital after the matched day. We also used a marginal structural Cox regression model^22^ to compare outcomes among patients who received dexamethasone plus remdesivir and patients who received remdesivir alone.

## RESULTS

### Patients

Of 2485 patients admitted to JHHS with COVID-19 between March 4 and August 29, 2020, 184 were excluded. Reasons for exclusion included death or discharge within 24 hours of admission (n=168), participation in randomized clinical trials of remdesivir (n=12), and being on remdesivir at the date of censoring (n=4). Among the remaining 2299 patients, 342 (14.9%) received remdesivir. Of these, 303 (88.6%) received a five-day course, 33 (9.6%) did not complete five days of treatment, and 6 (1.8%) received more than five days of treatment (**Figure 1**). The median time from admission to treatment initiation was 1.1 days (interquartile range [IQR] 0.8 – 2.5). After time-dependent propensity score matching, 303 (88.6%) patients in the remdesivir group were matched successfully and were selected for primary statistical analyses. **Table 1** shows the characteristics of remdesivir recipients and controls before (day 0) and after (day 1 of treatment) propensity score matching.

**Table 1.**
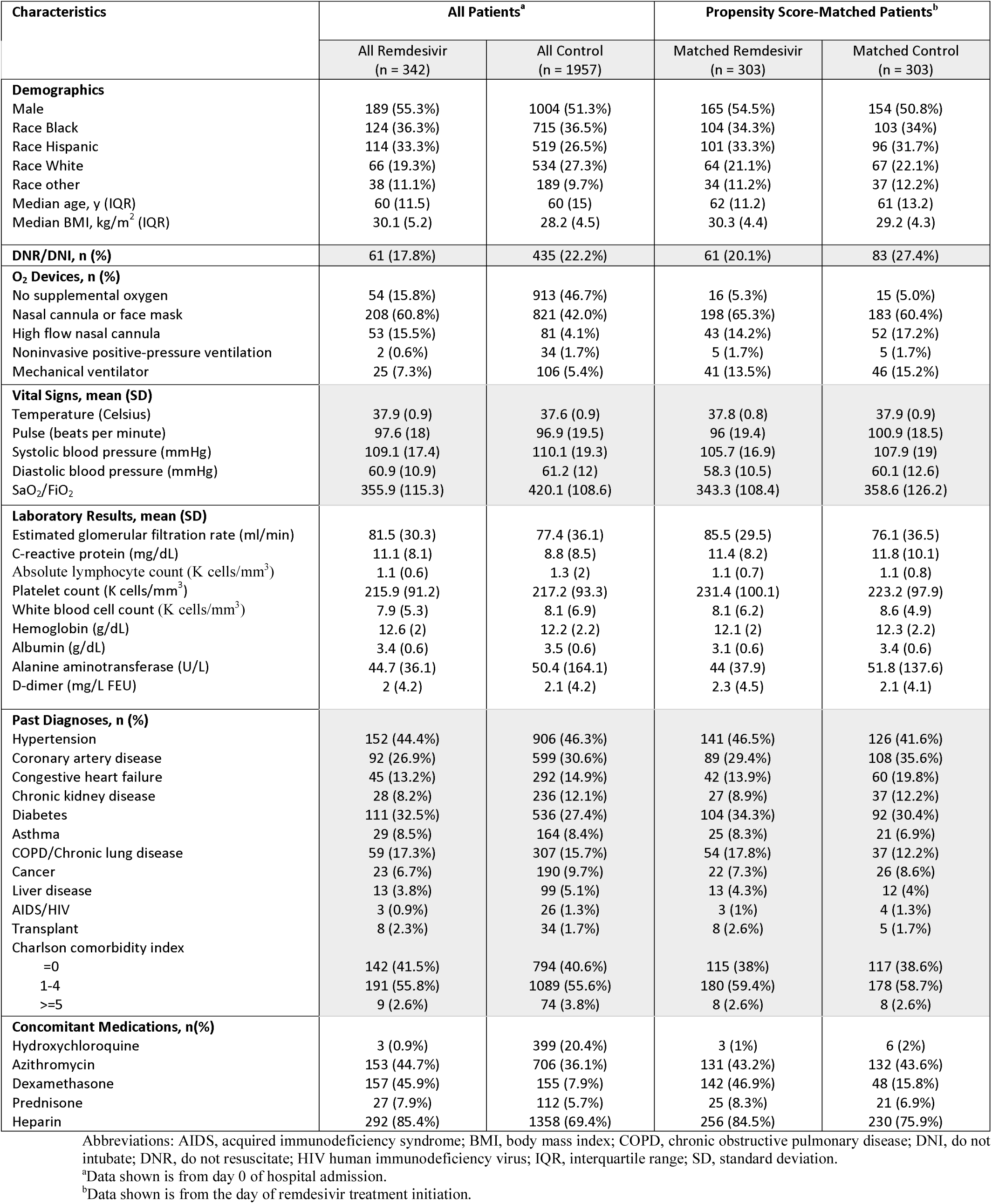
Characteristics of patients before and after propensity score matching

| Characteristics | All Patients<sup>a</sup> |  | Propensity Score-Matched Patients<sup>b</sup> |  |
| --- | --- | --- | --- | --- |
|  | All Remdesivir<br>(n = 342) | All Control<br>(n = 1957) | Matched Remdesivir<br>(n = 303) | Matched Control<br>(n = 303) |
| **Demographics** |  |  |  |  |
| Male | 189 (55.3%) | 1004 (51.3%) | 165 (54.5%) | 154 (50.8%) |
| Race Black | 124 (36.3%) | 715 (36.5%) | 104 (34.3%) | 103 (34%) |
| Race Hispanic | 114 (33.3%) | 519 (26.5%) | 101 (33.3%) | 96 (31.7%) |
| Race White | 66 (19.3%) | 534 (27.3%) | 64 (21.1%) | 67 (22.1%) |
| Race other | 38 (11.1%) | 189 (9.7%) | 34 (11.2%) | 37 (12.2%) |
| Median age, y (IQR) | 60 (11.5) | 60 (15) | 62 (11.2) | 61 (13.2) |
| Median BMI, kg/m<sup>2</sup> (IQR) | 30.1 (5.2) | 28.2 (4.5) | 30.3 (4.4) | 29.2 (4.3) |
| **DNR/DNI, n (%)** | 61 (17.8%) | 435 (22.2%) | 61 (20.1%) | 83 (27.4%) |
| **O<sub>2</sub> Devices, n (%)** |  |  |  |  |
| No supplemental oxygen | 54 (15.8%) | 913 (46.7%) | 16 (5.3%) | 15 (5.0%) |
| Nasal cannula or face mask | 208 (60.8%) | 821 (42.0%) | 198 (65.3%) | 183 (60.4%) |
| High flow nasal cannula | 53 (15.5%) | 81 (4.1%) | 43 (14.2%) | 52 (17.2%) |
| Noninvasive positive-pressure ventilation | 2 (0.6%) | 34 (1.7%) | 5 (1.7%) | 5 (1.7%) |
| Mechanical ventilator | 25 (7.3%) | 106 (5.4%) | 41 (13.5%) | 46 (15.2%) |
| **Vital Signs, mean (SD)** |  |  |  |  |
| Temperature (Celsius) | 37.9 (0.9) | 37.6 (0.9) | 37.8 (0.8) | 37.9 (0.9) |
| Pulse (beats per minute) | 97.6 (18) | 96.9 (19.5) | 96 (19.4) | 100.9 (18.5) |
| Systolic blood pressure (mmHg) | 109.1 (17.4) | 110.1 (19.3) | 105.7 (16.9) | 107.9 (19) |
| Diastolic blood pressure (mmHg) | 60.9 (10.9) | 61.2 (12) | 58.3 (10.5) | 60.1 (12.6) |
| SaO<sub>2</sub>/FiO<sub>2</sub> | 355.9 (115.3) | 420.1 (108.6) | 343.3 (108.4) | 358.6 (126.2) |
| **Laboratory Results, mean (SD)** |  |  |  |  |
| Estimated glomerular filtration rate (ml/min) | 81.5 (30.3) | 77.4 (36.1) | 85.5 (29.5) | 76.1 (36.5) |
| C-reactive protein (mg/dL) | 11.1 (8.1) | 8.8 (8.5) | 11.4 (8.2) | 11.8 (10.1) |
| Absolute lymphocyte count (K cells/mm<sup>3</sup>) | 1.1 (0.6) | 1.3 (2) | 1.1 (0.7) | 1.1 (0.8) |
| Platelet count (K cells/mm<sup>3</sup>) | 215.9 (91.2) | 217.2 (93.3) | 231.4 (100.1) | 223.2 (97.9) |
| White blood cell count (K cells/mm<sup>3</sup>) | 7.9 (5.3) | 8.1 (6.9) | 8.1 (6.2) | 8.6 (4.9) |
| Hemoglobin (g/dL) | 12.6 (2) | 12.2 (2.2) | 12.1 (2) | 12.3 (2.2) |
| Albumin (g/dL) | 3.4 (0.6) | 3.5 (0.6) | 3.1 (0.6) | 3.4 (0.6) |
| Alanine aminotransferase (U/L) | 44.7 (36.1) | 50.4 (164.1) | 44 (37.9) | 51.8 (137.6) |
| D-dimer (mg/L FEU) | 2 (4.2) | 2.1 (4.2) | 2.3 (4.5) | 2.1 (4.1) |
| **Past Diagnoses, n (%)** |  |  |  |  |
| Hypertension | 152 (44.4%) | 906 (46.3%) | 141 (46.5%) | 126 (41.6%) |
| Coronary artery disease | 92 (26.9%) | 599 (30.6%) | 89 (29.4%) | 108 (35.6%) |
| Congestive heart failure | 45 (13.2%) | 292 (14.9%) | 42 (13.9%) | 60 (19.8%) |
| Chronic kidney disease | 28 (8.2%) | 236 (12.1%) | 27 (8.9%) | 37 (12.2%) |
| Diabetes | 111 (32.5%) | 536 (27.4%) | 104 (34.3%) | 92 (30.4%) |
| Asthma | 29 (8.5%) | 164 (8.4%) | 25 (8.3%) | 21 (6.9%) |
| COPD/Chronic lung disease | 59 (17.3%) | 307 (15.7%) | 54 (17.8%) | 37 (12.2%) |
| Cancer | 23 (6.7%) | 190 (9.7%) | 22 (7.3%) | 26 (8.6%) |
| Liver disease | 13 (3.8%) | 99 (5.1%) | 13 (4.3%) | 12 (4%) |
| AIDS/HIV | 3 (0.9%) | 26 (1.3%) | 3 (1%) | 4 (1.3%) |
| Transplant | 8 (2.3%) | 34 (1.7%) | 8 (2.6%) | 5 (1.7%) |
| Charlson comorbidity index |  |  |  |  |
| =0 | 142 (41.5%) | 794 (40.6%) | 115 (38%) | 117 (38.6%) |
| 1-4 | 191 (55.8%) | 1089 (55.6%) | 180 (59.4%) | 178 (58.7%) |
| >=5 | 9 (2.6%) | 74 (3.8%) | 8 (2.6%) | 8 (2.6%) |
| **Concomitant Medications, n(%)** |  |  |  |  |
| Hydroxychloroquine | 3 (0.9%) | 399 (20.4%) | 3 (1%) | 6 (2%) |
| Azithromycin | 153 (44.7%) | 706 (36.1%) | 131 (43.2%) | 132 (43.6%) |
| Dexamethasone | 157 (45.9%) | 155 (7.9%) | 142 (46.9%) | 48 (15.8%) |
| Prednisone | 27 (7.9%) | 112 (5.7%) | 25 (8.3%) | 21 (6.9%) |
| Heparin | 292 (85.4%) | 1358 (69.4%) | 256 (84.5%) | 230 (75.9%) |

Abbreviations: AIDS, acquired immunodeficiency syndrome; BMI, body mass index; COPD, chronic obstructive pulmonary disease; DNI, do not intubate; DNR, do not resuscitate; HIV human immunodeficiency virus; IQR, interquartile range; SD, standard deviation.
<sup>a</sup>Data shown is from day 0 of hospital admission.
<sup>b</sup>Data shown is from the day of remdesivir treatment initiation.

**Figure 1.**
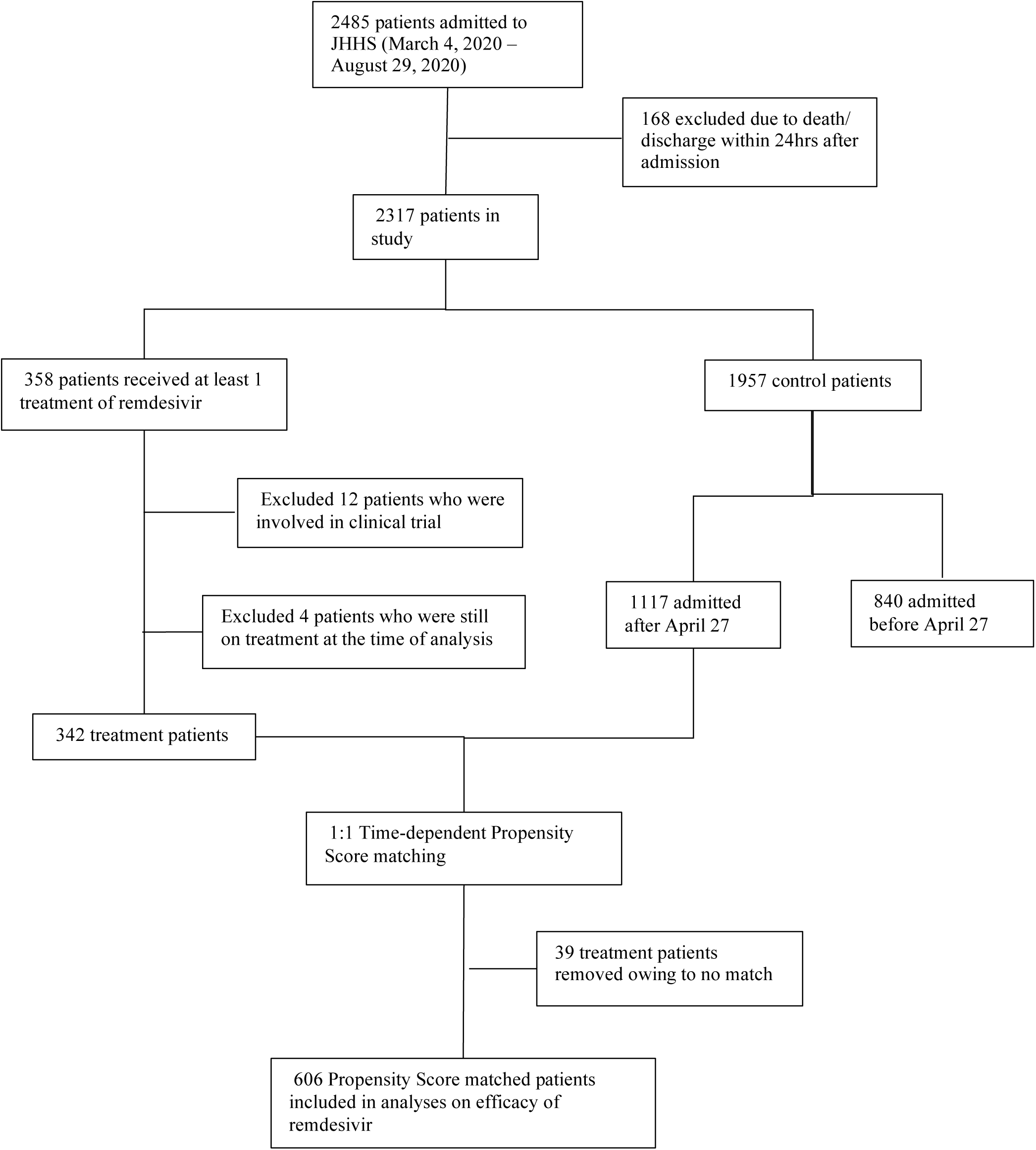
Description of patients included in the analysis.

### Primary Outcome

Among a total of 606 matched individuals (303 remdesivir and 303 matched controls), 251 (82.8%) remdesivir patients and 223 (73.6%) controls achieved clinical improvement before 28 days with a median time to clinical improvement of 5.0 days (IQR 4.0 – 8.0) and 7.0 days (IQR 5.0 – 12.0 days), respectively. In Cox proportional hazards models, remdesivir significantly shortened time to clinical improvement (aHR 1.55 [1.28; 1.87]) (**Figure 2a**). Remdesivir patients on room air or nasal cannula oxygen had a significantly more rapid time to clinical improvement compared to matched controls (median 5 days [IQR 4.0 – 7.0] vs. median 6 days [IQR 5.0 – 10.0]; aHR 1.54 [1.22;1.93]); those with severe disease (requiring higher levels of respiratory support) did not significantly benefit from remdesivir (median 8.0 days [IQR 6.0 – 13.0], remdesivir vs. median 11.0 days [IQR 6.0 –15.5], control; aHR 1.39 [0.91; 2.11]) (**Figure 2b-c**).

**Figure 2.**
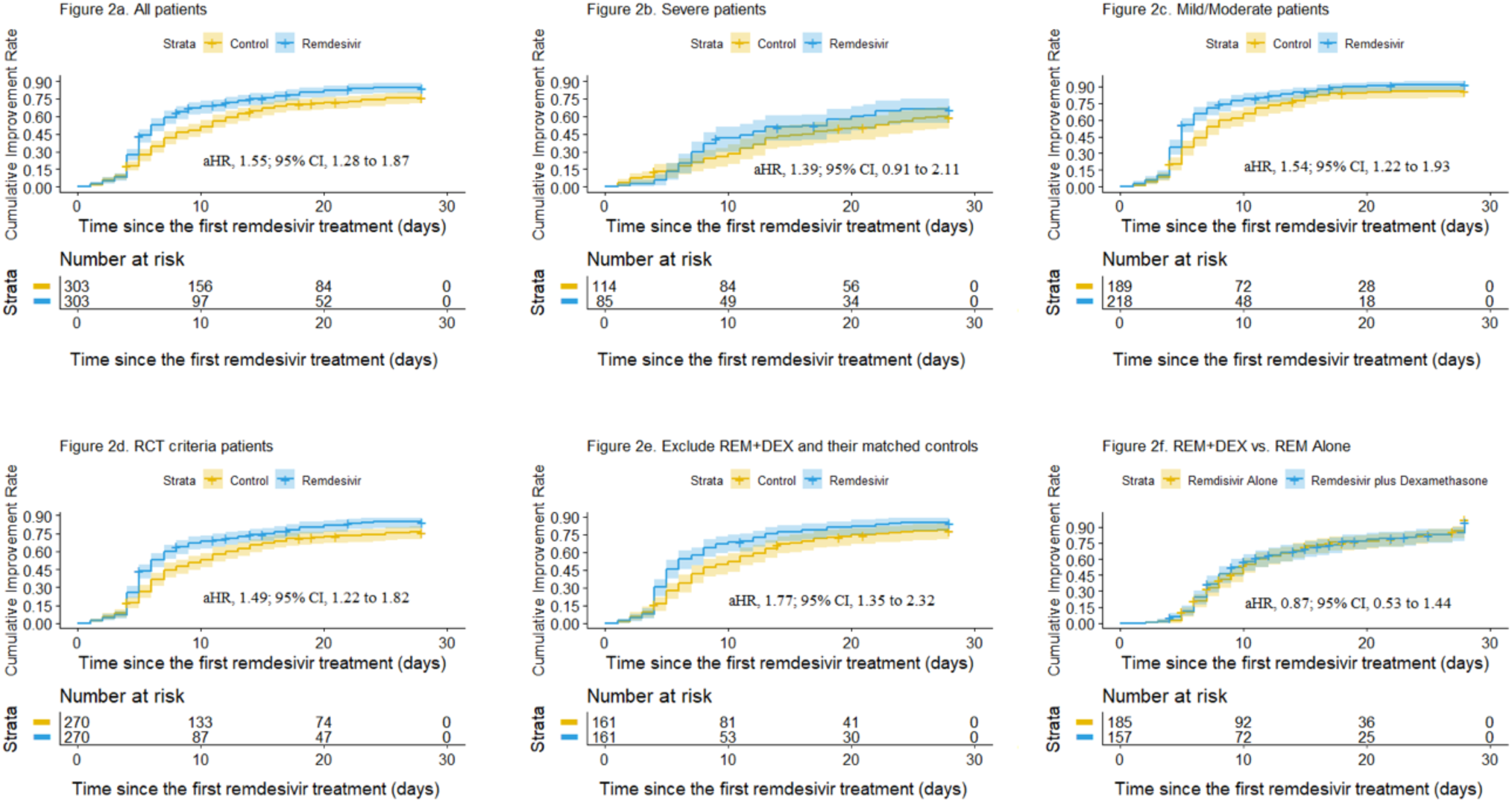
Cumulative incidence curves for time to clinical improvement are shown for: the entire remdesivir and matched-control cohort (Panel A); patients with severe disease (requiring high-flow nasal cannula, non-invasive ventilation, mechanical ventilation, ECMO or vasopressors) (Panel B); patients with mild to moderate disease (on room air or nasal cannula oxygen) (Panel C); patients who would have met ACTT-1 study criteria (Panel D); patients who did not receive dexamethasone (Panel E); and patients who received remdesivir plus dexamethasone compared to patients who received remdesivir alone (Panel F).

We performed a secondary analysis considering only the 306 remdesivir patients who met inclusion criteria for ACTT-1 (**eFigure1)**. Of these, 270 (88.2%) were successfully matched with control patients who also satisfied the criteria (**eTable1**). The median time to clinical improvement was 5.0 days (IQR 4.0 – 8.0) among remdesivir patients and 7.0 days (IQR 5.0 – 11.0) among matched controls; remdesivir significantly shortened time to clinical improvement in Cox regression analysis (aHR 1.49 [1.22 – 1.82]) (**Figure 2d**).

### Secondary Outcome

Remdesivir recipients had a 28-day mortality of 7.6% (23 deaths) compared to 14.9% (45 deaths) among matched controls, and median time to death was 8.4 days (IQR 6.2 – 12.8) and 9.5 days (IQR 6.7 – 13.8), respectively. All discharges (257 remdesivir and 225 controls) were treated as censored at 28 days. Remdesivir did not have a statistically significant impact on time to death (aHR 0.80 [0.46; 1.41]) (**Figure 3a**) and offered no significant mortality benefit among patients in the mild to moderate disease group (aHR 0.78 [0.27; 2.28]) or the severe disease group (aHR 0.94 [0.43; 2.03]) (**Figure 3b-c**). When we restricted the analysis to patients who would have met the inclusion criteria for ACTT-1, 28-day mortality did not change appreciably (6.3%, remdesivir vs. 14.1%, control). Median time to death was 9.0 days (IQR 7.1 – 17.4) and 8.5 days (IQR 6.7 – 13.6) among remdesivir and control patients, respectively, and remdesivir did not have a significant impact on time to death in cox regression analysis (aHR 0.55 [0.29; 1.06]) (**Figure 3d**).

**Figure 3.**
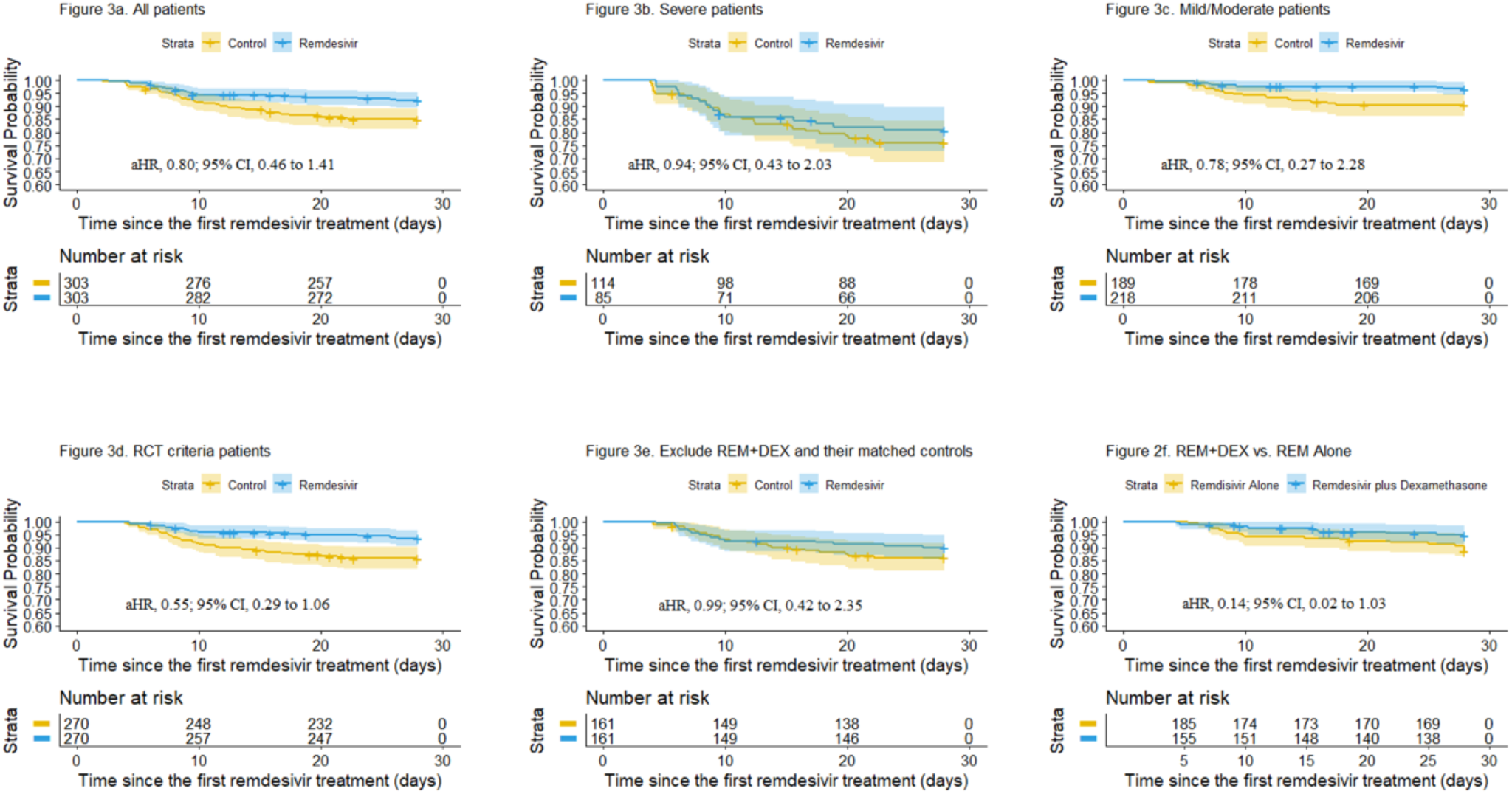
Kaplan-Meier survival curves are shown for: the entire remdesivir and matched-control cohort (Panel A); patients with severe disease (requiring high-flow nasal cannula, non-invasive ventilation, mechanical ventilation, ECMO or vasopressors) (Panel B); patients with mild to moderate disease (on room air or nasal cannula oxygen) (Panel C); patients who would have met ACTT-1 study criteria (Panel D); patients who did not receive dexamethasone (Panel E); and patients who received dexamethasone plus remdesivir compared to patients who received remdesivir alone (Panel F).

**Figure 4.**
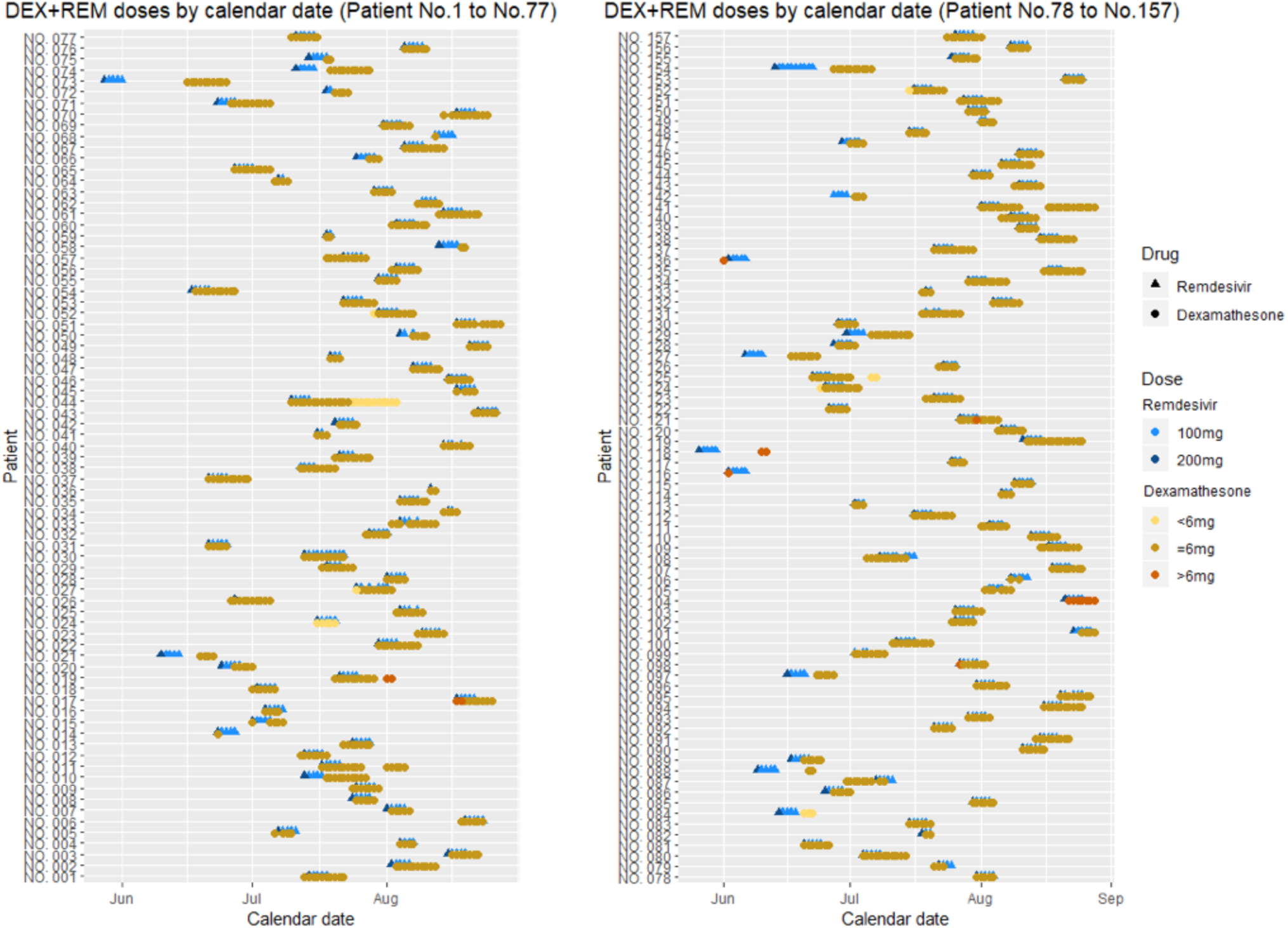
The time-course for co-administration of remdesivir and dexamethasone is shown for the 157 patients who received both dexamethasone and remdesivir. The majority of patients received 6mg of dexamethasone daily as utilized in the RECOVERY trial.

### Sensitivity Analyses

To account for the potential impact of dexamethasone, we excluded 157 patients who received remdesivir plus dexamethasone (and their matched controls) from our initial analyses. Remdesivir alone still had a significant impact on time to clinical improvement (aHR 1.77 [1.35; 2.32]) (**Figure 2e**) and no significant effect on mortality (aHR 0.99 [0.42; 2.35]) (**Figure 3e**). These results remained unchanged when we excluded patients who received any type of corticosteroid treatment (e.g. dexamethasone, prednisone, prednisolone, methylprednisolone or hydrocortisone) (data not shown).

We also compared patients who received remdesivir plus dexamethasone (n=157) to patients who received remdesivir alone (n=185) (**eTable2**). Combination therapy had a more rapid time to clinical improvement (7.0 days (IQR 5.0 – 12.0], combination vs. 8.0 days (IQR 6.0 – 14.0], control), but did not have a significant effect in a marginal structural Cox regression model (aHR 0.87 [0.53; 1.44]) (**Figure 2f**). 28-day mortality was also lower in the combination therapy group (5.1% [8 deaths], combination vs 9.2% [17 deaths], control) (**eTable2**). A marginal structural Cox regression model showed a trend towards reduced time to death with remdesivir plus dexamethasone (aHR 0.14 [0.02; 1.03]) (**Figure 3f**).

The results are sensitive to the requirement that controls be selected from among patients who remained hospitalized during the same period of treatment as their matched counterpart (up to 5 days). Although there is no qualitative change if the requirement is reduced to 4 days, the estimated aHR decreases to 1.0 [0.2;1.25] if the required period of hospitalization is lowered to 3 days or less. This is because 90 patients in the control group had their event within 4 days of matching compared to 25 patients in the treatment group.

### Reasons for Stopping Remdesivir Early

Of the 33 patients (9.6%) who did not complete at least 5 days of remdesivir, 21 were discharged and 2 died before completing treatment. Among the remaining patients, treatment was stopped for the following reasons: liver enzyme or bilirubin elevation (n=4), renal failure of unclear etiology (n=2), nausea (n=1), epistaxis and tachycardia (n=1), neck and mouth itching (n=1) and transition to comfort care (n=1). The incidence of liver enzymes above 200 IU, bilirubin above 2 mg/dL as well as estimated glomerular filtration rate less than 30 ml/min is shown in (**eTable3)**.

## DISCUSSION

Optimal implementation of therapeutics to reduce COVID-19 morbidity and mortality is a global priority. In this retrospective multicenter study, individuals who received remdesivir had a statistically significant reduction in time to clinical improvement of 2 days, but no statistically significant decrease in mortality. Patients with mild disease were more likely to benefit compared to those with severe disease. The magnitude and direction of these effects were similar to the benefits seen in ACTT-1.^8^ Our study shows that remdesivir is beneficial in populations that have been underrepresented in clinical trials (i.e. Black and Latinx individuals) and supports a 5-day treatment course. There was also an intriguing trend towards reduced mortality among patients who received remdesivir plus dexamethasone, indicating that this combination of antiviral and anti-inflammatory therapy warrants further study.

Our study included a much higher percentage of underrepresented minorities than previous remdesivir clinical trials. Approximately 90% of patients in our cohort were non-white compared to 30-47% in clinical trials of remdesivir.^8,10,11^ Since underrepresented minorities have shouldered a disproportionate burden during COVID-19, but have not been widely represented in clinical trials,^12^ our results provide important evidence that the benefits of remdesivir on time to clinical improvement apply to these populations.

Notably, the vast majority of patients in the remdesivir group received 5-days of therapy, which supports recommendations for an initial 5-day course for most patients. While the ACTT-1 trial used a 10-day treatment course, a comparison of 5 days versus 10 days showed similar efficacy^10^, and a recent open-label study found that a 5-day course, but not a 10-day course, was associated with a significant improvement in disease severity.^11^ Our finding that a 5-day treatment course is sufficient to see a clinical benefit is important, particularly given reports of both U.S. and global shortages of remdesivir and the potential for even greater supply constraints in the months to come.^23^

Our cohort included several groups who were not eligible to receive remdesivir in prior clinical trials, such as individuals who were pregnant (9 patients), under the age of 18 (2 patients) and who had evidence of chronic kidney disease at baseline (16 patients). Inclusion of these patients did not lead to an increased incidence of significant side effects as evidenced by the small percentage of patients in our cohort who stopped treatment early (2.9%) compared to the ACTT-1 trial (9%).

Finally, there was a trend towards improved mortality in patients who received both remdesivir and dexamethasone. This supports the hypothesis that an antiviral combined with an anti-inflammatory might improve outcomes in COVID-19.^24^ Our study as well as ACTT-1 showed that patients on room air or nasal cannula oxygen benefitted from remdesivir, while patients receiving higher levels of respiratory support, such as mechanical ventilation, did not benefit.^8^ The Food and Drug Administration (FDA) recently revised the remdesivir EUA to remove the requirement that patients must be hypoxemic or require supplemental oxygen.^25^ In contrast, the RECOVERY trial showed that only patients receiving supplemental oxygen or additional respiratory support benefitted from dexamethasone whereas patients on room air did not.^16^ The combination of dexamethasone and remdesivir, the appropriate target population and the timing of co-administration warrant additional study.

Our study has limitations. The time-dependent propensity score methodology produced matched sets in which patients were very similar on measured confounders. However, there could be unmeasured variables that biased our treatment effect estimates. It is possible there was a secular trend in the quality of COVID-19 patient care as our health system gained experience, so we limited our matched control group to the time period when remdesivir was available. We required controls to remain hospitalized for the period of time remdesivir patients were on treatment, not exceeding 5 days. This was a pre-analysis design to avoid bias from matching controls with different disease severity that would not be controlled for by the measurement confounders. However, our findings are sensitive to this restriction being reduced to 3 or fewer days. This is consistent with the ACTT-1 trial that excluded patients expected to be discharged within 3 days of randomization.^8^ We also took into account the increased use of dexamethasone after the RECOVERY trial.^16^ Removing patients who received remdesivir plus dexamethasone (or corticosteroids in general) did not appreciably change the overall findings.

In conclusion, remdesivir use in patients who met the EUA criteria led to a significant decrease in the time to clinical recovery in patients admitted to the hospital for COVID-19. These results provide further evidence that remdesivir is effective in reducing the duration of COVID-19 illness, that a 5-day treatment course may be sufficient and that patients with milder disease benefit most. The inclusion of a larger proportion of underrepresented minorities provides much needed evidence of effectiveness in these groups. Additional studies to assess the clinical benefit of combination therapy with remdesivir and dexamethasone in COVID-19 infected patients are warranted.

## CONTRIBUTORS

BTG, KW, YX contributed to the conception and design of the work, the acquisition, analysis, and interpretation of data,

MR, SZ, KBR, MCW, CA, AG, RB contributed to the conception and design of the work and interpretation of the data.

All authors contributed to the drafting and revising of the manuscript for important intellectual content, give final approval of the version to be published and agree to be accountable for all aspects of the work in ensuring that questions related to the accuracy or integrity of any part of the work are appropriately investigated and resolved.

## Data Availability

Available by request

## ACKNOWLEDGMENTS

The data utilized for this publication were part of the JH-CROWN: The COVID PMAP Registry, which is based on the contribution of many patients and clinicians. The authors would like to thank the Johns Hopkins Health System and its surrounding communities for working together to provide outstanding patient care and to keep each other safe during these extraordinary times.

## FUNDING

This work was supported by funding from Hopkins inHealth, the Johns Hopkins Precision Medicine Program through JH-CROWN and the COVID-19 Administrative Supplement for the HHS Region 3 Treatment Center from the Office of the Assistant Secretary for Preparedness and Response (to BTG, KW, MR, AG and YX).

## CONFLICTS OF INTEREST

Dr. Alexander is past Chair of the Food and Drug Administration Peripheral and Central Nervous System Advisory Committee; has served as a paid advisor to IQVIA; is a co-founding Principal and equity holder in Monument Analytics, a health care consultancy whose clients include the life sciences industry as well as plaintiffs in opioid litigation; and is a member of OptumRx’s National P&T Committee. These arrangements have been reviewed and approved by Johns Hopkins University in accordance with its conflict of interest policies.

## Notes

### Competing Interest Statement

All authors have completed the ICMJE uniform disclosure form and declare: Dr Alexander has served as a paid advisor to IQVIA, is a cofounding Principal and equity holder in Monument Analytics, is a member of the OptumRx National P and T Committee, and former Chair of the Food and Drug Administration Peripheral and Central Nervous System Advisory Committee. These arrangements have been reviewed and approved by Johns Hopkins University in accordance with its conflict of interest policies.

### Author Declarations

The Institutional Review Boards at Johns Hopkins Hospital, Baltimore, MD; Bayview Hospital, Baltimore, MD; Howard County General Hospital, Columbia, MD; Suburban Hospital, Bethesda, MD; Sibley Hospital, Washington DC, approved this study as minimal risk and waived informed consent requirements.

