## Supplemental Materials for "Effectiveness of remdesivir with and without dexamethasone in hospitalized patients with COVID-19"

**Supplementary Appendix**

This appendix has been provided by the authors to give readers additional information about their work.

### **List of Investigators**

**Brian T. Garibaldi MD MEPH**^*^ - Division of Pulmonary and Critical Care, Johns Hopkins University School of Medicine, Baltimore MD

**Kunbo Wang MS^*^**- Department of Applied Mathematics and Statistics, Johns Hopkins University, Baltimore, MD

**Matt Robinson MD** - Division of Infectious Disease, Johns Hopkins University School of Medicine, Baltimore, MD

**Scott Zeger PhD** - Division of Biostatistics, Johns Hopkins Bloomberg School of Public Health

**Karen Bandeen Roche PhD** - Division of Biostatistics, Johns Hopkins Bloomberg School of Public Health

**Mei-Cheng Wang PhD** - Division of Biostatistics, Johns Hopkins Bloomberg School of Public Health

**G. Caleb Alexander MD** - Center for Drug Safety and Effectiveness, Johns Hopkins Bloomberg School of Public Health, Baltimore, MD

**Amita Gupta MD** - Division of Infectious Disease, Johns Hopkins University School of Medicine

**Robert Bollinger MD MPH** - Division of Infectious Disease, Johns Hopkins University School of Medicine

**Yanxun Xu PhD** - Department of Applied Mathematics and Statistics, Johns Hopkins University, Baltimore, MD; Division of Biostatistics and Bioinformatics at The Sidney Kimmel Comprehensive Cancer Center, Johns Hopkins University School of Medicine, Baltimore, MD

* Dr. Garibaldi and Mr. Wang contributed equally to this article.

### **eMethods**

#### **Time-dependent Propensity Score Matching**

Since the timing of initial remdesivir administration was different and the remdesivir assignment was non-randomized, a time-dependent propensity score matching method was applied to undertake 1:1 propensity score matching for the treatment arm so that additional analyses could be performed on the matched patients. Propensity scores were calculated from a time-dependent Cox regression model using the time to the first receipt of remdesivir as the outcome, where the propensity score at a given hospitalization day is the hazard of exposure to remdesivir treatment at that day.^1-2^

Time-dependent covariates in records before the first remdesivir administration date (for treatment group patients) and records before right-censoring or last follow-up date (for control group patients) were used as predictors to obtain parameter estimates. Time-invariant (fixed) covariates included race, age, sex, body mass index (BMI), Charlson comorbidity index (CCI), and code status (i.e. whether the patient had a “do not resuscitate” order). Time-dependent (varying) covariates included various clinical measures of disease severity, such as SaO2/FiO2 ratio, systolic blood pressure (SBP), diastolic blood pressure (DBP), pulse, temperature, respiratory rate, and supplemental Oxygen device (O2 device). SaO2/FiO2 ratio was selected since it can be calculated without obtaining an arterial blood gas (not available on all patients). Other time-dependent variables included key laboratory test results, such as C-reactive protein (CRP), absolute lymphocyte count (ALC), Platelets count (PLC), White blood cell count (WBC), Hemoglobin (HGB), Albumin, Alanine aminotransferase (ALT), Estimated Glomerular Filtration Rate (eGFR), and D-dimer.

Beginning from day zero, a sequential 1:1 greedy matching without replacement was conducted. Patients were included in the matching process only if their admission dates were later than the earliest admission date (April 27, 2020) of patients in the remdesivir group. A patient who first received remdesivir at a given day *t* of hospitalization was matched with those who did not, based on their propensity scores (hazard components) at day *t*. In addition, a time constraint was imposed so that a patient in the remdesivir group with *k* days of treatment, was forced to match a patient in the control group who stayed at least *k* days (5 days maximum) in the hospital since the matched day. This constraint removed patients from the control group who were healthy enough to be discharged in one or two days from the matched day as those patients were unlikely to receive remdesivir treatment at the matched day if they were close to discharge.

#### **Marginal Structural Cox Regression Model**

In sub-analyses considering the effects of combination drug use, comparisons of patients who used combination of dexamethasone and remdesivir (n=157) with patients who used remdesivir alone (n=185) were conducted. Since the sample sizes for two groups were similar, and patients’ exposure to dexamethasone was time-variant, instead of time-dependent propensity score matching, Marginal Structural Cox regression models were conducted to adjust for the non-randomized administration of dexamethasone, in the meantime, to analyze the effects of dexamethasone on outcomes of interests in patients who had exposure to remdesivir.^3^ The same set of time-dependent covariates and time-invariant variables as in the matching models from all remdesivir patients were included in the model. Inverse Probability Treatment Weighting (IPTW) method was applied for parameter estimation.

#### **Outcome of Interest Analyses**

The primary outcome of interests was time to clinical improvement from the treatment start of remdesivir, defined as discharge alive from the hospital without worsening of their WHO disease severity score or at least a two-point decrease in the WHO severity score during hospitalization within 28 days or max follow-up after the first treatment of remdesivir. Failure of clinical improvement was censored at max follow-up day or 28 days, whichever came first. Death was also censored at 28 days. The secondary outcome was time to death from the first remdesivir treatment day. Patients who were discharged alive were censored at 28 days.

Cox proportional-hazards regression models were applied to estimate the association between remdesivir treatment and outcomes of interests. A set of demographics, clinical variables and laboratory test results were included in Cox regression models based on clinical interest and knowledge. Time-invariant variables included race, age, sex, body mass index (BMI), Charlson comorbidity index (CCI), and code. Time dependent covariates included SaO2/FiO2 ratio, SBP, DBP, CRP, ALC, Respiratory rate, Temperature, Pulse, WBC, HGB, Albumin, ALT, eGFR, and D-dimer. The adjusted hazard ratio of remdesivir treatment was estimated from Cox regression model after controlling these covariates.

#### **Missing Data Imputations**

For the laboratory results, missing values were imputed using the last observation carried forward if the last observation was within three days of the missing data, otherwise, using multiple imputation by chained equations (MICE) with predictive mean matching method.^4^

**eFigure1.** Exclusion Details under ACTT-1 Criteria

342 remdesivir treatment patients

306 patients satisfied ACTT-1 criteria

36 were excluded:

- 2 due to age < 18yrs

- 9 due to pregnancy

- 1 due to PaO_2_≥ 94% on room air

- 16 due to eGFR < 30

- 8 due to ALT or AST > 200

270 patients were matched successfully

**eFigure1 Legend.** ACTT-1 Criteria: patients were excluded if age < 18 years, or pregnant, or PaO_2_ ≥ 94% on room air, or either alanine aminotransferase (ALT) or aspartate aminotransferase (AST) > 200, or estimated glomerular filtration rate (eGFR) < 30.

**eTable1.** Characteristics of Patients Satisfied ACTT-1 Criteria, before and after Propensity-Score Matching

| Characteristics | *All Remdesivir Patients*  *Satisfied Criteria*^†^  *(n = 306)* | Propensity Score – Matched Patients^†^ | |
| --- | --- | --- | --- |
|  |  | *Matched Remdesivir (n=270)* | *Matched Control*  *(n=270)* |
| **Demographics:**  Male  Race Black  Race Hispanic  Race White  Race Others  Age, Median (IQR)  BMI, Median (IQR) | 176 (57.5%)  105 (34.3%)  103 (33.7%)  62 (20.3%)  36 (11.8%)  60.5 (10)  31.1 (5.1) | 156 (57.8%)  87 (32.2%)  91 (33.7%)  58 (21.5%)  34 (12.6%)  62 (10.4)  30.2 (4.4) | 145 (53.7%)  97 (35.9%)  85 (31.5%)  54 (20%)  34 (12.6%)  61 (12.4)  29.6 (4.5) |
| **DNR/DNI, no. (%)** | 53 (17.3%) | 53 (19.6%) | 72 (26.7%) |
| **O2 Devices, no. (%):**  No Supplemental Oxygen  Nasal Cannula or Face Mask  High Flow Nasal Cannula  Noninvasive Positive-Pressure Ventilation  Mechanical Ventilator | 11 (3.6%)  193 (63.1%)  54 (17.6%)  4 (1.3%)  44 (14.4%) | 11 (4.1%)  180 (66.7%)  37 (13.7%)  4 (1.5%)  38 (14.1%) | 45 (16.7%)  146 (54.1%)  35 (13.0%)  9 (3.3%)  33 (12.2%) |
| **Vital Signs, Mean (SD):**  Temperature (^o^Celcius)  Pulse (beats per minute)  Systolic BP (mmHg)  Diastolic BP (mmHg)  SaO_2_/FiO_2_ | 37.9 (0.9)  96 (18.4)  105.6 (16.7)  58.2 (10.5)  334.4 (110.4) | 37.8 (0.8)  96 (18.7)  106 (16.9)  58.6 (10.6)  341.8 (109.4) | 37.9 (0.9)  102 (18.2)  107.3 (18)  60.2 (12.1)  357.5 (126.2) |
| **Laboratory Results, Mean (SD):**  Estimated glomerular filtration rate (ml/min)  C-reactive protein (mg/dL)  Absolute lymphocyte count (K cells/mm^3^)  Platelets count (K cells/mm^3^)  White blood cell count (K cells/mm^3^)  Hemoglobin (g/dL)  Albumin (g/dL)  Alanine aminotransferase (U/L)  D-dimer (mg/L FEU) | 89.9 (27.2)  11.7 (8.2)  1.1 (0.7)  236.7 (107.4)  8.1 (6.1)  12.2 (1.9)  3.1 (0.6)  43.6 (31.8)  2.2 (4.5) | 87.9 (25.9)  11.6 (8.3)  1.1 (0.6)  234.1 (102.5)  8 (6.3)  12.2 (1.9)  3.2 (0.6)  43.3 (31.9)  2.2 (4.3) | 82.7 (32.4)  12.1 (10.2)  1.1 (1.1)  223.6 (100)  8.6 (4.8)  12.5 (2.2)  3.4 (0.6)  47.7 (105.9)  2.1 (4.3) |
| **Past Diagnoses, no. (%)**  Hypertension  Coronary Artery Disease  Congestive heart failure  Chronic kidney disease  Diabetes  Asthma  COPD/Chronic Lung Disease  Cancer  Liver Disease  AIDS/HIV  Transplant  Charlson Comorbidity Index:  =0  1-4  >=5 | 131 (42.8%)  83 (27.1%)  38 (12.4%)  17 (5.6%)  96 (31.4%)  24 (7.8%)  53 (17.3%)  20 (6.5%)  11 (3.6%)  3 (1%)  7 (2.3%)  128 (41.8%)  172 (56.2%)  6 (2%) | 119 (44.1%)  79 (29.3%)  36 (13.3%)  17 (6.3%)  88 (32.6%)  20 (7.4%)  48 (17.8%)  19 (7%)  11 (4.1%)  3 (1.1%)  7 (2.6%)  104 (38.5%)  161 (59.6%)  5 (1.9%) | 102 (37.8%)  85 (31.5%)  47 (17.4%)  15 (5.6%)  77 (28.5%)  21 (7.8%)  35 (13%)  24 (8.9%)  8 (3%)  2 (0.7%)  4 (1.5%)  115 (42.6%)  149 (55.2%)  6 (2.2%) |
| **Concomitant Medications, no. (%):**  Hydroxychloroquine  Azithromycin  Dexamethasone  Prednisone  Heparin | 1 (0.3%)  137 (44.8%)  143 (46.7%)  22 (7.2%)  269 (87.9%) | 1 (0.4%)  117 (43.3%)  130 (48.1%)  20 (7.4%)  235 (87%) | 6 (2.2%)  120 (44.4%)  40 (14.8%)  19 (7%)  226 (83.7%) |

† Data shown is from the day of remdesivir treatment initiation

**eTable2.** Characteristics of Remdesivir Patients Stratified by Dexamethasone Use, before and after Matching

| Characteristics | All Remdesivir Patients^†^ | |
| --- | --- | --- |
|  | Rem Alone (n = 185) | Rem + Dex (n = 157) |
| **Demographics:**  Male  Race Black  Race Hispanic  Race White  Race Others  Age, Median (IQR)  BMI, Median (IQR) | 101 (54.6%)  66 (35.7%)  75 (40.5%)  28 (15.1%)  16 (8.6%)  57 (12.5)  31.2 (5.4) | 88 (56.1%)  58 (36.9%)  39 (24.8%)  38 (24.2%)  22 (14%)  63 (9.5)  30.9 (3.5) |
| **DNR/DNI, no. (%)** | 32 (17.3%) | 29 (18.5%) |
| **O2 Devices, no. (%):**  No Supplemental Oxygen  Nasal Cannula or Face Mask High Flow Nasal Cannula  Noninvasive Positive-Pressure Ventilation  Mechanical Ventilator | 2 (4.4%)  20 (44.5%)  13 (28.9%)  1 (2.2%)  9 (20.0%) | 8 (4.5%)  108 (61.4%)  29 (16.5%)  2 (1.1%)  29 (16.5%) |
| **Vital Signs, Mean (SD):**  Temperature (^o^Celcius)  Pulse (beats per minute)  Systolic BP (mmHg)  Diastolic BP (mmHg)  SaO_2_/FiO_2_ | 38 (0.8)  98.9 (19.7)  104.5 (16.6)  57.5 (9.7)  338.3 (113.1) | 37.7 (0.9)  93.6 (17.6)  106.7 (17.3)  58.6 (11.4)  330.3 (106) |
| **Laboratory Results, Mean (SD):**  Estimated glomerular filtration rate (ml/min)  C-reactive protein (mg/dL)  Absolute lymphocyte count (K cells/mm^3^)  Platelets count (K cells/mm^3^)  White blood cell count (K cells/mm^3^)  Hemoglobin (g/dL)  Albumin (g/dL)  Alanine aminotransferase (U/L)  D-dimer (mg/L FEU) | 89.3 (32.3)  12.4 (8.7)  1.1 (0.6)  237.2 (110.2)  8.1 (7.2)  11.9 (1.9)  3 (0.6)  44.4 (39)  2.9 (5.5) | 84.7 (29.5)  10.7 (8)  1 (0.7)  233 (99.4)  8 (4)  12.2 (2.1)  3.3 (0.6)  44.8 (35)  1.6 (3.3) |
| **Past Diagnoses, no. (%)**  Hypertension  Coronary Artery Disease  Congestive heart failure  Chronic kidney disease  Diabetes  Asthma  COPD/Chronic Lung Disease  Cancer  Liver Disease  AIDS/HIV  Transplant  Charlson Comorbidity Index:  =0  1-4  >=5 | 78 (42.2%)  48 (25.9%)  28 (15.1%)  15 (8.1%)  61 (33%)  14 (7.6%)  26 (14.1%)  15 (8.1%)  6 (3.2%)  2 (1.1%)  3 (1.6%)  81 (43.8%)  99 (53.5%)  5 (2.7%) | 74 (47.1%)  44 (28%)  17 (10.8%)  13 (8.3%)  50 (31.8%)  15 (9.6%)  33 (21%)  8 (5.1%)  7 (4.5%)  1 (0.6%)  5 (3.2%)  61 (38.9%)  92 (58.6%)  4 (2.5%) |
| **Concomitant Medications, no. (%):**  Hydroxychloroquine  Azithromycin  Prednisone  Heparin | 3 (1.6%)  88 (47.6%)  15 (8.1%)  160 (86.5%) | 0 (0%)  65 (41.4%)  12 (7.6%)  132 (84.1%) |

† Data shown is from the day of remdesivir treatment initiation

Rem denotes Remdesivir

Dex denotes Dexamethasone

**eTable3.** Adverse Events

|  | Matched Remdesivir  (n = 303) | Matched Control  (n = 303) |
| --- | --- | --- |
| **Before Matched Day or Drug Day, no. (%):**  ALT or AST > 200 IU  bilirubin > 2 mg/dL  eGFR < 30 ml/min | 5 (1.7%)  3 (1.0%)  3 (1.0%) | 7 (2.3%)  7 (2.3%)  7 (2.3%) |
| **On/After Matched Day or Drug Day, no. (%):**  ALT or AST > 200 IU  bilirubin > 2 mg/dL  eGFR < 30 ml/min | 31 (10.2%)  11 (3.6%)  27 (8.9%) | 29 (9.6%)  18 (5.9%)  63 (20.8%) |
| **Anytime in hospital, no. (%):**  ALT or AST > 200 IU  bilirubin > 2 mg/dL  eGFR < 30 ml/min | 34 (11.2%)  12 (4.0%)  32 (10.6%) | 34 (11.2%)  22 (7.3%)  77 (25.4%) |
